# Bayesian nowcasting with Laplacian-P-splines

**DOI:** 10.1101/2022.08.26.22279249

**Authors:** Bryan Sumalinab, Oswaldo Gressani, Niel Hens, Christel Faes

## Abstract

During an epidemic, the daily number of reported infected cases, deaths or hospitalizations is often lower than the actual number due to reporting delays. Nowcasting aims to estimate the cases that have not yet been reported and combine it with the already reported cases to obtain an estimate of the daily cases. In this paper, we present a fast and flexible Bayesian approach to do nowcasting by combining P-splines and Laplace approximations. The main benefit of Laplacian-P-splines (LPS) is the flexibility and faster computation time compared to Markov chain Monte Carlo (MCMC) algorithms that are often used for Bayesian inference. In addition, it is natural to quantify the prediction uncertainty with LPS in the Bayesian framework, and hence prediction intervals are easily obtained. Model performance is assessed through simulations and the method is applied to COVID-19 mortality and incidence cases in Belgium.

## 1 Introduction

Nowcasting is a term used for estimating the occurred-but-not-yet-reported-events (Donker et al., 2011; Van de Kassteele et al., 2019). In epidemiology, real-time updates of new symptomatic/infected individuals are helpful to assess the present situation and provide recommendations for rapid planning and for implementing essential measures to contain an epidemic outbreak. The exact number of new daily cases is frequently subject to reporting delays, resulting in underreporting of the real number of infected individuals for that day. Failing to account for the reporting delays will lead to possibly biased predictions that might have an effect on policy making (Gutierrez et al., 2020). The main goal of nowcasting is to estimate the actual number of new cases by combining the (predicted) not-yet-reported-cases with the already reported cases.

Several early references that establish the statistical framework for this type of problem can be found in the paper of Lawless (1994); De Angelis and Gilks (1994); and Lind-sey (1996). In disease surveillance, Höhle and an der Heiden (2014) applied nowcasting to the outbreak of Shiga toxin-producing Escherichia coli in Germany and also to the SARS-CoV-2 outbreak (Glöckner et al., 2020; Günther et al., 2021). Their approach is formulated within a hierarchical Bayesian framework that consists of estimating the epidemic curve by using a quadratic spline based on a truncated power basis function and the time-varying reporting delay distribution which is approximated by a discrete time survival model. Van de Kassteele et al. (2019) pointed out that a potential drawback of such a method is the long computation time required by the Markov chain Monte Carlo (MCMC) algorithm and therefore proposed an alternative fast and flexible modelling strategy based on bivariate P-splines (Penalized B-splines). P-splines (Eilers and Marx, 1996) provide a flexible smoothing tool used to describe trends in the data. It introduces a penalty parameter that controls the roughness of the fit and counterbalances the flexibility of a rich B-splines basis (Eilers and Marx, 1996; Eilers et al., 2015). Other attractive features of the P-splines smoother are the relatively simple structure of the penalty matrix that is effortlessly computed and the natural extension to a Bayesian framework (Lang and Brezger, 2004). Based on the approach of Van de Kassteele et al. (2019), the number of cases are structured in a two-dimensional table (with calendar time as the first dimension and delay time as the second dimension), yielding the data matrix used as an input in the model. The reporting intensity is assumed to be a smooth surface and is modelled using two-dimensional P-splines.

In this paper, we build upon the work of Van de Kassteele et al. (2019) by proposing a new nowcasting methodology based on Laplacian-P-splines (LPS) in a fully Bayesian framework. A key advantage of working with the Bayesian approach is the ease to obtain the predictive distribution and quantify the uncertainty associated with the predictions. In addition, the posterior distribution of the penalty parameter can be explored, and hence its uncertainty can be accounted for. The Laplace approximation uses a second-order Taylor expansion to approximate the posterior distribution of the regression parameters by a Gaussian density. It is a sampling-free method with the major advantage of faster computational time as opposed to MCMC approaches that are commonly used in Bayesian inference. Therefore, given the flexibility of previously mentioned P-splines smoothers and the computational benefit of Laplace approximations, it can be a helpful tool in the daily monitoring of new cases during an epidemic period. Laplacian-P-splines already proved to be useful in survival models (Gressani and Lambert, 2018; Gressani et al., 2022a), generalized additive models (Gressani and Lambert, 2021) and also in epidemic models for estimating the effective reproduction number (Gressani et al., 2022b). We build on the work of Gressani and Lambert (2021) to extend the Laplacian-P-splines methodol-ogy to nowcasting, thereby providing a fast and flexible (fully) Bayesian alternative to Van de Kassteele et al. (2019). To evaluate the (predictive) performance of our method, a simulation study is implemented and several performance measures are reported such as the (relative) bias, prediction interval coverage, and prediction interval width. Finally, we apply our method to the COVID-19 mortality and incidence data in Belgium for the year 2021 and 2022, respectively. The R codes used for the simulation study are available on GitHub through the link https://github.com/bryansumalinab/Laplacian-P-spline-nowcasting.git.

## 2 Methodology

### 2.1 Bayesian model formulation

In this section, we follow the work of Van de Kassteele et al. (2019) to formulate a fully Bayesian model based on P-splines. Let *y*_*t*,*d*_ denote the number of cases that occurred at time *t* = 1, 2, …, *T* (corresponding to the calendar day) and are reported with a delay of *d* = 0, 1, 2…, *D* days. The information on cases can be summarized in matrix form:

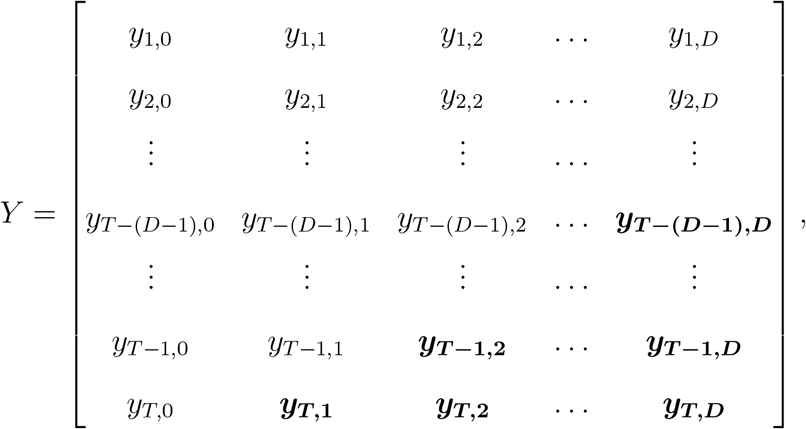

with cases that have not yet been reported (at time *T* ) highlighted in bold. The not-yet-reported cases correspond to (*t, d*) combinations satisfying *t* > *T* − *d*. The main objective is to predict the total number of cases, 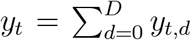, for *t* = *T* − (*D* − 1),..., *T* for which the nowcasted and already reported cases can be combined.

Let 𝒟:= ***y*** = (*y*_1_, *y*_2_ … , *y*_*n*_)^*⊤*^ denote the vector of the observed number of cases by stacking the columns of matrix *Y* for the reported cases, where each entry corresponds to each (*t, d*) combination of reported cases *y*_*t*,*d*_. The model assumes that the number of cases either follows a Poisson or a negative binomial (NB) distribution, i.e., *y*_*t*,*d*_ ∼ Poisson(*µ*_*t*,*d*_) or *y*_*t*,*d*_ ∼ NB(*µ*_*t*,*d*_, *ϕ*) with mean *µ*_*t*,*d*_ *>* 0. For the negative binomial, *ϕ >* 0 is an overdispersion parameter and the variance is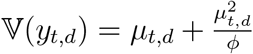. Following Van de Kassteele et al. (2019), the (log) mean number of cases is modeled with two dimensional B-splines:

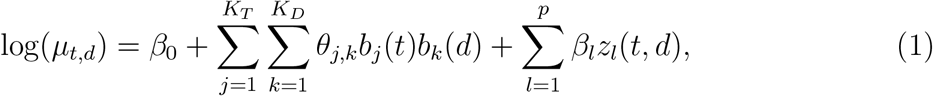

where *β*_0_ is the intercept; *b*_*j*_(·) and *b*_*k*_(·) are univariate B-splines basis functions specified in the time and delay dimensions, respectively; and *z*_*l*_(*t, d*) represents additional covariates with regression coefficients *β*_*l*_. In matrix notation:

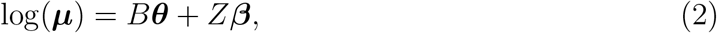

where the matrices *B* and *Z* correspond to the basis functions and covariates, respectively, and vectors ***θ*** and ***β*** are the associated parameters to be estimated (Details in supplementary material S1).

In the philosophy of P-splines (Eilers and Marx, 1996), we use a rich (cubic) B-splines basis and counterbalance the associated flexibility by imposing a discrete roughness penalty on contiguous B-spline coefficients. For the two dimensional P-splines, the penalty can be obtained based on row-wise (direction of calendar time) and column-wise (direction of reporting delay) differences for matrix Θ = (*θ*_*j*,*k*_) with *j* = 1, … , *K*_*T*_ and *k* = 1, … , *K*_*D*_ (supplementary material S1) (see Durbán et al. (2002) and Fahrmeir et al. (2013) (pp. 507-508)). Let 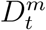 and 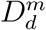 denote the *m*th order row-wise and column-wise difference matrix with dimensions (*K*_*T*_ − *m*) × *K*_*T*_ and (*K*_*D*_ − *m*) × *K*_*D*_, respectively. In this paper, we use a second order (*m* = 2) difference penalty. For ease of notation, let 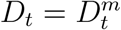 and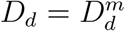. The difference matrix for vector ***θ*** can be obtained by expanding the difference matrix into 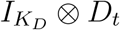 and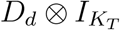 , where ⊗ denotes the Kronecker product. Using this notation, the row-wise and column-wise difference penalty can be written, respectively, as:

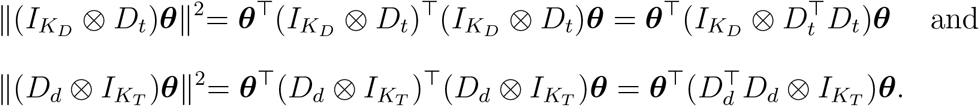

Let *λ*_*t*_ *>* 0 and *λ*_*d*_ *>* 0 denote the row-wise and column-wise penalty parameter, respectively. The penalty for the two dimensional B-splines is then given by:

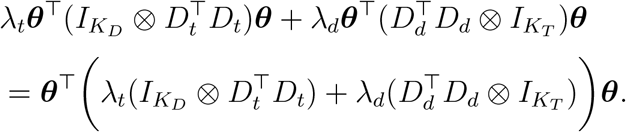

Let us define the penalty matrices 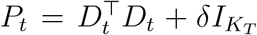 and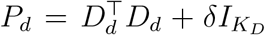, where *δ* is a small number (say *δ* = 10^−12^), to ensure that the penalty matrices are full rank and thus invertible. Following Lang and Brezger (2004), the penalty can be translated in the Bayesian framework by specifying a Gaussian prior (***θ***|***λ***) ∼ 𝒩_*dim*(***θ***)_(**0**, *𝒫*^−1^(***λ***)), where **λ** = (λ_*t*_, λ_*d*_)^⊤^ is the penalty vector and 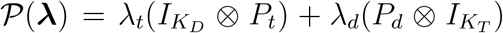 is the penalty matrix. A Gaussian prior is also assumed for ***β***, namely 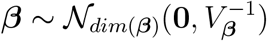 with *V*_***β***_ = *ζI*_*p*+1_ (small *ζ*, e.g. *ζ* = 10^−5^). Denote by *X* = (*B, Z*) the global design matrix, ***ξ*** = (***β***^*⊤*^, ***θ***^*⊤*^)^*⊤*^ the latent parameter vector and 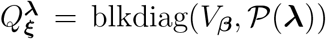 the precision matrix for ***ξ*** where blkdiag(·) refers to a block diagonal matrix. From here, we focus on the negative binomial model for the number of cases. The Poisson model is described in detail in supplementary material S4. The full Bayesian (negative binomial) model is summarized as follows:

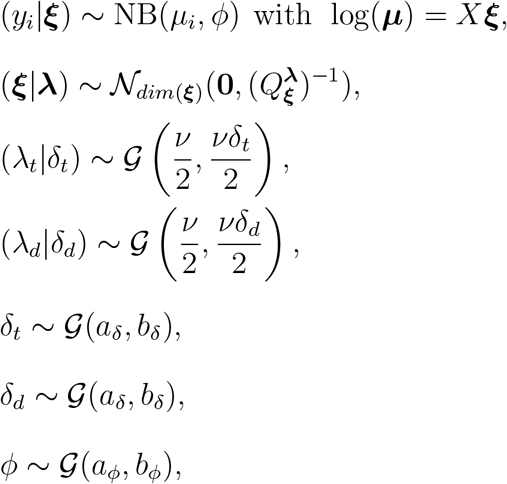

where 𝒢 (*a, b*) denotes a Gamma distribution with mean *a/b* and variance *a/b*^2^. This robust prior specification on the penalty parameters follows from Jullion and Lambert (2007). They have shown that when the hyperparameters *a*_*δ*_, *b*_*δ*_ are chosen to be equal and small enough (say 10^−5^), then the resulting fit is robust to the value of *ν* (e.g. *ν* = 3 in this paper). In addition, we also set *a*_*ϕ*_ = *b*_*ϕ*_ = 10^−5^.

### 2.2 Laplace approximation to the conditional posterior of *ξ*

The conditional posterior of the latent vector ***ξ*** is approximated by a Gaussian distribution via the Laplace approximation. The gradient and Hessian of the (log) conditional posterior are analytically derived and used in a Newton-Raphson algorithm to obtain the Gaussian approximation to the conditional posterior distribution of ***ξ***.

Note that for a negative binomial distributed *y*_*i*_ with mean *E*(*y*_*i*_) = *µ*_*i*_, the probability distribution can be written as an exponential dispersion family given by *p*(*y*_*i*_; *γ, ϕ*) = exp{[*y*_*i*_*γ*_*i*_ − *b*(*γ*_*i*_)]*/a*(*ϕ*) + *c*(*y*_*i*_, *ϕ*)}, where 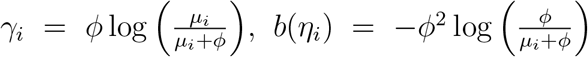, 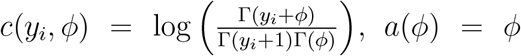 and Γ(·) is the gamma function. Thus, for fixed *ϕ*, a negative binomial regression model is a generalized linear model (GLM) where *µ*_*i*_ is linked on the linear predictor through the link function *g*(*µ*_*i*_) such that 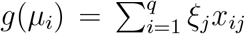 (see Agresti (2013) for a detailed account about GLMs). Here, we use the log-link function *g*(*µ*_*i*_) = log(*µ*_*i*_). The log-likelihood is given by log 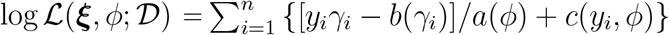 and it can be shown that for a negative binomial model, the gradient and Hessian are:

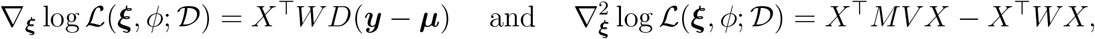

where *W* = diag(*w*_1_, … , *w*_*n*_), *w*_*i*_ = [𝕍 (*y*_*i*_)(*g*′(*µ*_*i*_))^2^]^−1^, *D* = diag(*g*′(*µ*_1_), … , *g*′(*µ*_*n*_)), *V* = diag(*υ*_1_, *υ*_2_, … , *υ*_*n*_) 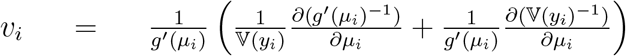 and *M* = diag(*y*_1_ − *µ*_1_, … , *y*_*n*_ − *µ*_*n*_) (see details in supplementary material S2).

Using Bayes’ rule, the posterior of ***ξ*** conditional on the penalty vector ***λ*** and overdispersion parameter *ϕ* is:

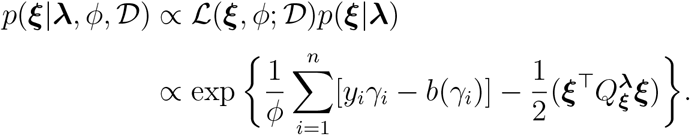

The gradient and Hessian for the log-conditional posterior of ***ξ*** are:

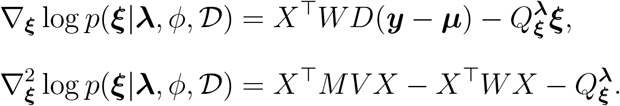

The above gradient and Hessian can then be used in a Newton-Raphson algorithm to obtain the mode of the conditional posterior of ***ξ***. After convergence, the Laplace approximation of the conditional posterior of ***ξ*** is a multivariate Gaussian density denoted by 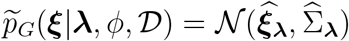 where 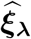 is the mean/mode and 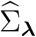 is the covariance matrix. There are instances where this approximation may be insufficient, for example, when dealing with sparse likelihoods or low counts. In such scenarios, the resulting posterior distribution could exhibit skewness. Lambert and Gressani (2023) proposed an approach to address this asymmetry issue but due to complexity, this is beyond the scope of this paper.

### 2.3 Optimization of hyperparameters and overdispersion parameter

In this section, we derive the (approximate) posterior distribution of the hyperparameters ***λ*** = (*λ*_*t*_, *λ*_*d*_)^⊤^ and ***δ*** = (*δ*_*t*_, *δ*_*d*_)^⊤^, and overdispersion parameter *ϕ*. Let ***η*** = (*λ*_*t*_, *λ*_*d*_, *δ*_*t*_, *δ*_*d*_)^⊤^ denote the vector of hyperparameters. Using Bayes’ theorem, the joint marginal posterior of ***η*** and *ϕ* is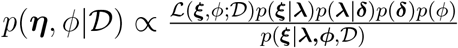. Following Rue et al. (2009), the above posterior can be approximated by replacing *p*(***ξ***|***λ***, *ϕ, 𝒟*) with 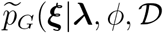 obtained in Section 2.2 and by evaluating the latent vector at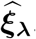. Note that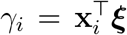, where 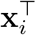 corresponds to the *i* th row of the design matrix *X*. Also, the determinant 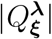 in *p*(***ξ***|***λ***) is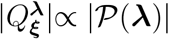. Hence, the approximated joint marginal posterior of ***η*** and *ϕ* is:

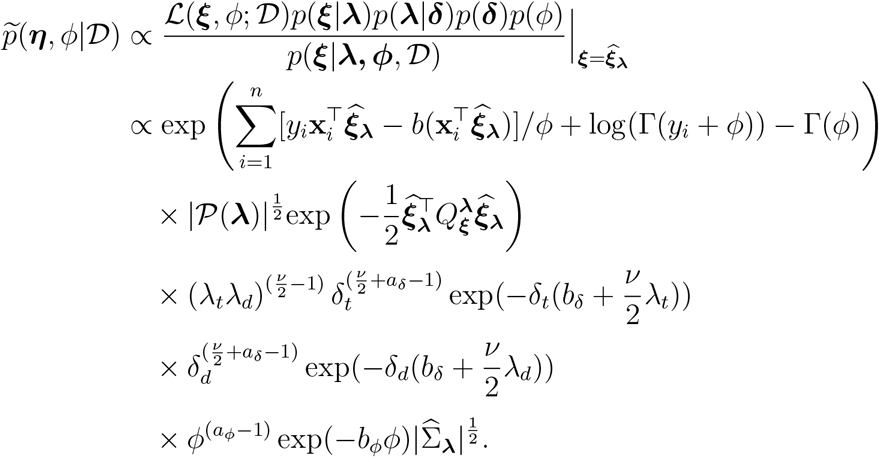

Integrating out the hyperparameters *δ*_*t*_ and *δ*_*d*_ from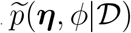, we obtain the joint marginal posterior of ***λ*** and *ϕ* as:

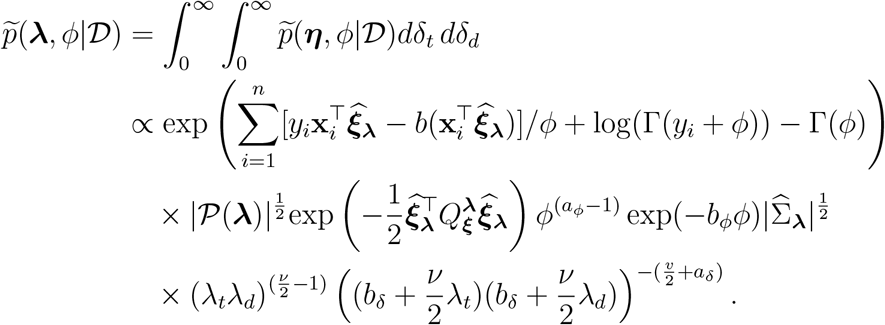

To ensure numerical stability, we log-transform the penalty vector ***υ*** = (*υ*_*t*_, *υ*_*d*_)^*⊤*^ = (log(*λ*_*t*_), log(*λ*_*d*_))^*⊤*^ and overdispersion parameter *υ*_*ϕ*_ = log(*ϕ*). The joint log-posterior of ***υ*** and *υ*_*ϕ*_ is then given by:

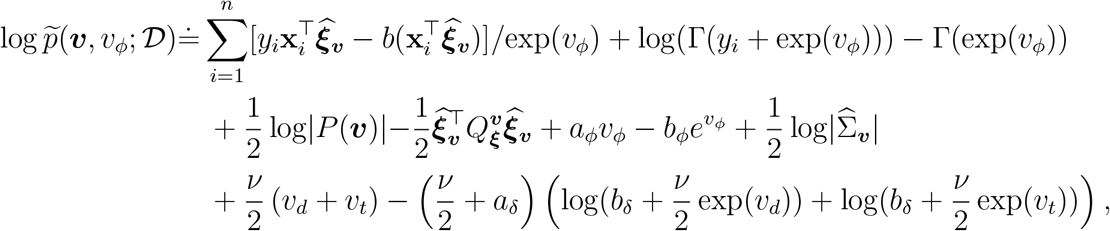

where ≐ denotes equality up to an additive constant. Finally, we maximize log 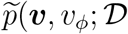 using the *optim()* function in R to obtain the posterior mode as a point estimate for ***v*** and *v*_*ϕ*_.

### 2.4 Nowcasting with prediction interval

To obtain the mean nowcast estimate with the prediction interval, note that Log 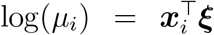 or equivalently log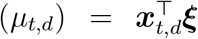, where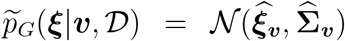. Thus,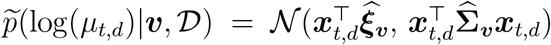. The mean estimate for the not-yet-reported cases is calculated as 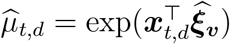 for all (*t, d*) combinations with *t* > *T* −*d*. Then, the estimate for the total number of cases for each *t* is obtained by summing the already reported cases and the mean estimate for the not-yet-reported cases, i.e.

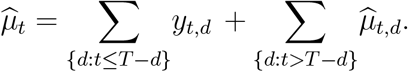

The prediction interval for the nowcast values is obtained by sampling from the posterior predictive distribution of the log-mean number of cases by following a five-step procedure:

1. For each (*t, d*) combinations with *t* > *T* − *d* (corresponding to the not-yet-reported cases), generate 1000 random samples 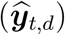 from a Gaussian distribution with mean 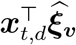 and variance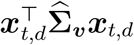.
2. Exponentiate the sampled values from the previous step to obtain the average reporting intensities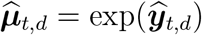.
3. Compute the average prediction for the not-yet-reported cases for each *t*. That is, compute 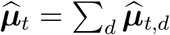 for *t* > *T* − *d*.
4. For each *t*, sample 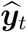 containing 1000 values from NB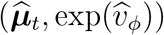.
5. Finally, compute the quantiles of the sampled values 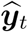 that correspond desired prediction interval.

### 2.5 Delay distribution

To obtain the smooth estimate of the delay distribution, only the first term on the right hand side of equation (2) is used (excluding the day of the week effects), as explained in the paper of Van de Kassteele et al. (2019). Specifically, the procedure to compute the delay distribution is as follows:

1. Compute the contribution of the smoothing term in equation (2) to the reporting intensity for all (*t, d*) combinations: ***µ***^smooth^ = exp(*B****θ***).
2. Arrange ***µ***^smooth^ into a *T ×* (*D* + 1) matrix with entries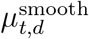.
3. For each *t* = 1, … , *T* , compute the reporting delay distribution given by: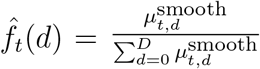

## 3. Simulations

A simulation study is implemented in order to evaluate the predictive performance of the proposed method. The procedure to perform the simulations is as follows:

1. Consider a function *f* (*t*) that represents the mean epidemic curve of all cases such that *µ*(*t*) = exp(*f* (*t*)) for *t* = 1, …, *T* .
2. For each *t*, generate a random sample *y*_*t*_ from a negative binomial distribution with mean *µ*(*t*) and fixed overdispersion parameter (we choose the value of 10).
3. To account for possible delays *d* = 0, 1, 2, … , *D*, generate samples from a multinomial distribution with probabilities (*p*_0_, *p*_1_, *p*_2_, *· · ·* , *p*_*D*_ such that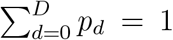), i.e.,

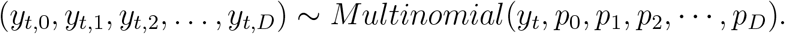

1. This sample represents the reported number of cases for each (*t, d*) combination. Repeat steps 1 to 3 for 500 times to generate 500 possible realizations.

We consider two functions inspired from the paper of Noufaily et al. (2016) given by:

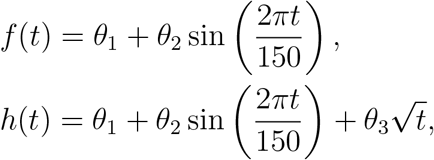

for *t* = 1, … , 365. That is, we assume a one year (365 days) time window in the simulation. In terms of delay probabilities, we consider a maximum delay of *D* = 7 days with probabilities (*p*_0_, *p*_1_, *p*_2_, *· · ·* , *p*_7_) = (0.0, 0.1, 0.4, 0.2, 0.1, 0.1, 0.05, 0.05), as illustrated in Figure 1. For example, 5% of the cases that occurred at time *T* − 6 have a delay of 7 days (with probability *p*_7_ = 0.05) and still need to be reported, or equivalently, 95% of the cases that occurred at time *T* − 6 have already been reported. For cases that occurred at time *T* − 5, only 90% of these cases have been reported, as cases with delays of 6 and 7 days are yet to be reported. For those cases that occurred on the nowcast day (time *T* ), none of these cases is reported, that is, the delay is 100%.

**Figure 1.**
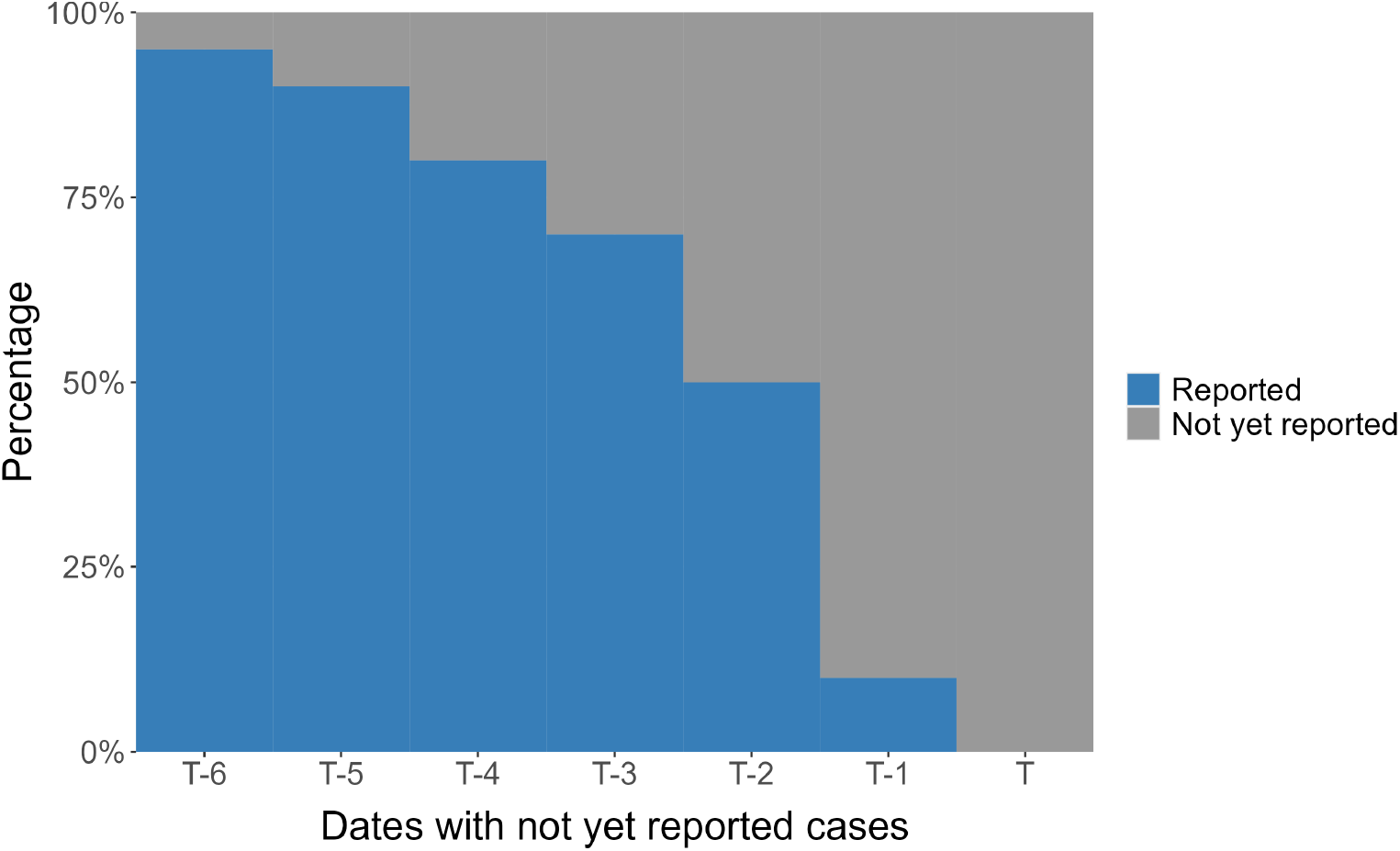
Illustration of the delay probabilities considered in the simulation.

Two scenarios are considered for *f* (*t*): (i) the first has a small number of cases with values of *θ*_1_ = 3 and *θ*_2_ = 1; (ii) the second scenario has a relatively large number of cases with *θ*_1_ = 3 and *θ*_2_ = 2 and an additional factor of 50 is added to the mean function such that *µ*(*t*) = 50 + exp(*f* (*t*)). These scenarios are denoted by *f*_1_(*t*) and *f*_2_(*t*), respectively. Similarly, two scenarios are also considered for the second function *h*(*t*): (i) the first scenario has values *θ*_1_ = 0, *θ*_2_ = 0.4 and *θ*_3_ = 0.2; (ii) the second scenario has values *θ*_1_ = 1.5, *θ*_2_ = 0.4 and *θ*_3_ = 0.2. We denote these functions by *h*_1_(*t*) and *h*_2_(*t*), respectively. The first function is symmetric with three peaks as shown in Figures 2a and 2b. On the other hand, the second function is not periodic as opposed to the first function (see Figures 2c and 2d). The plots for (one realization of) simulated cases based on these functions are shown in Figure 3 with (dashed) vertical lines corresponding to the different nowcast dates.

**Figure 2.**
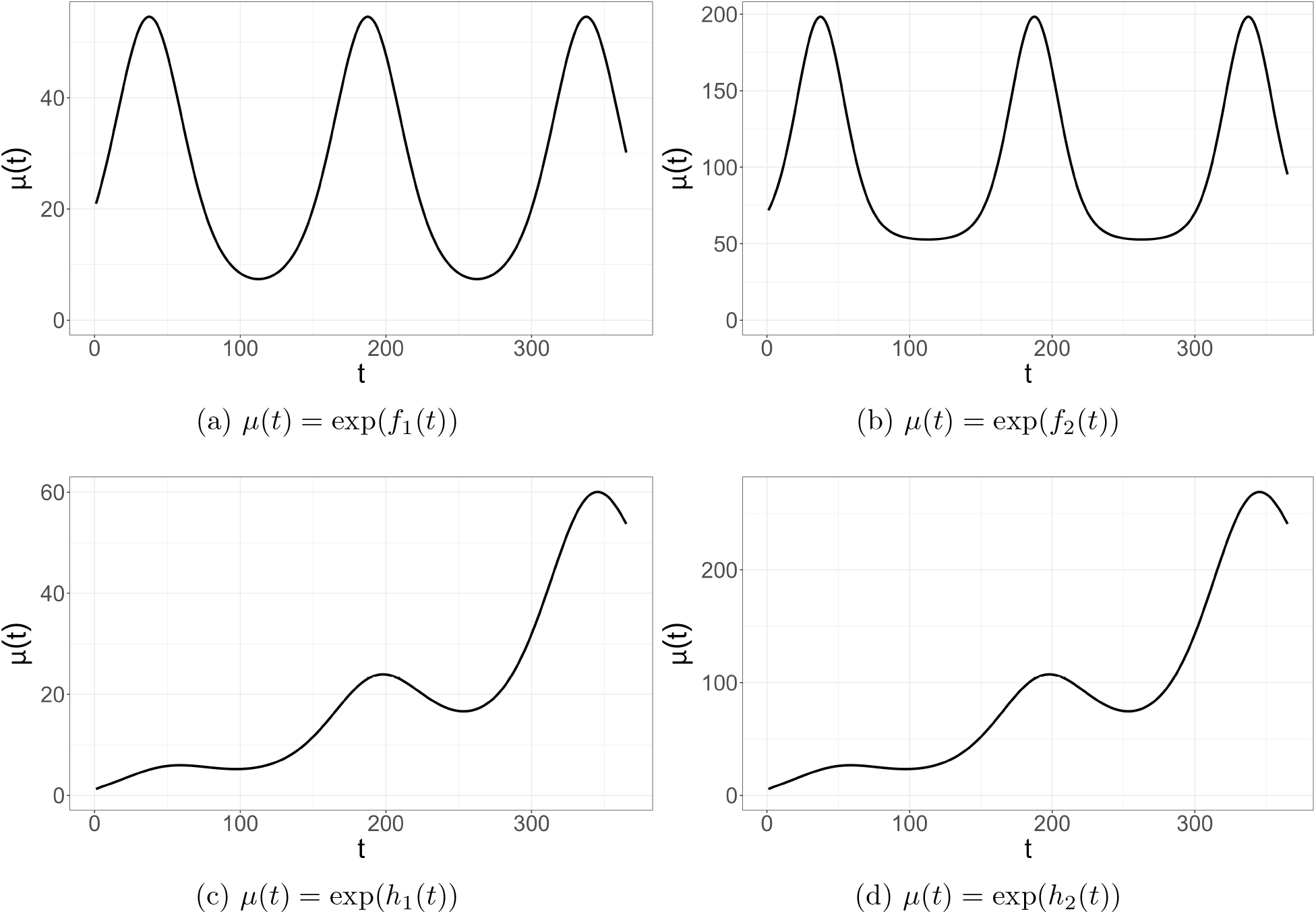
Mean epidemic curves considered in the simulations.

**Figure 3.**
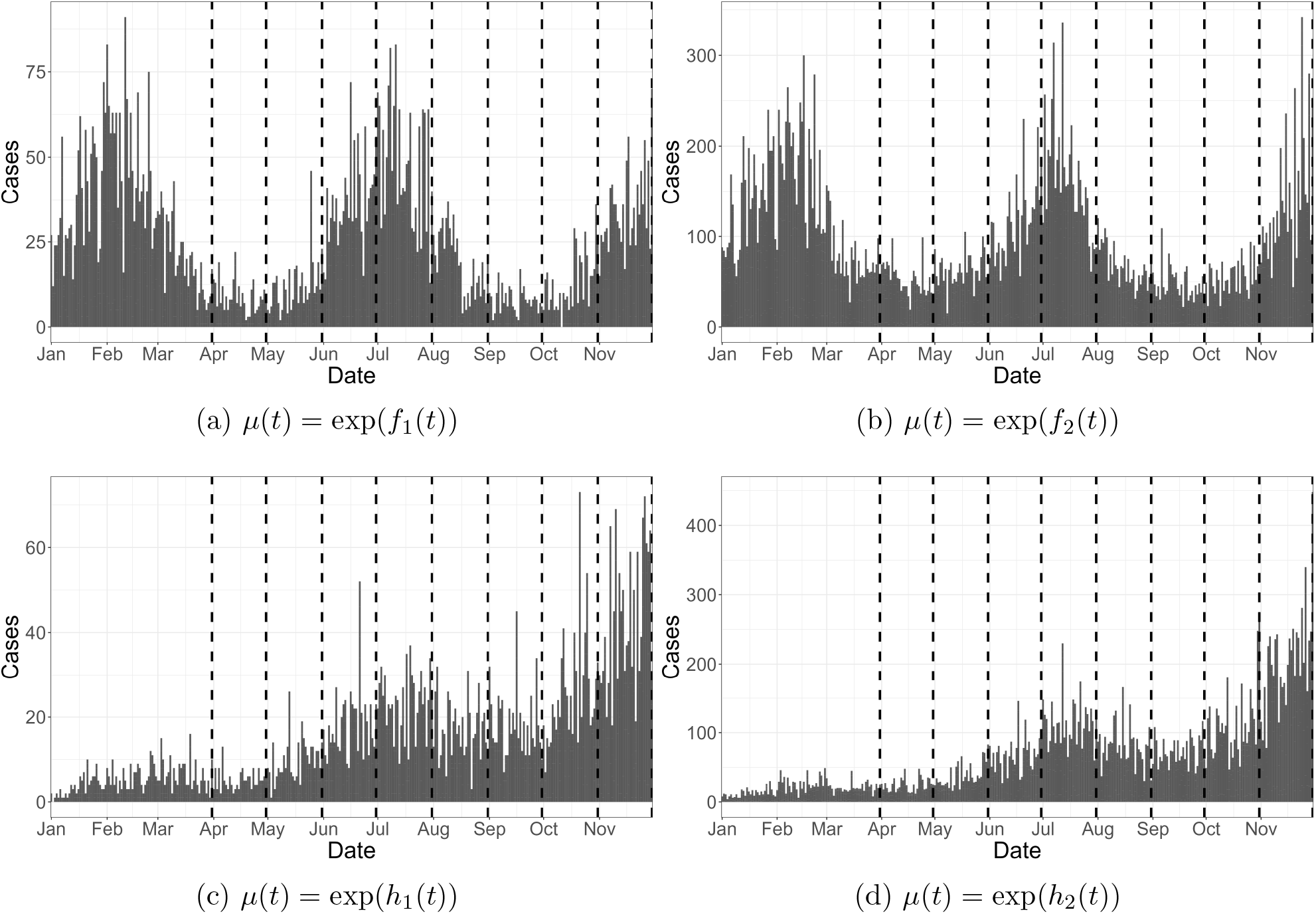
Simulated cases for different epidemic curves. The dashed vertical lines correspond to different nowcast dates.

For each generated data set, we fit the proposed models (LPS-NB and LPS-Poisson) as well as the method proposed by Van de Kassteele et al. (2019) (VDK) on the data, and compute the desired accuracy measures. The bias and relative bias (% bias) for the true mean epidemic curve (***µ***), 95% prediction interval (PI) coverage and prediction interval (PI) width are chosen to measure the predictive accuracy of our methodology. For a given calendar day *t*, the bias and relative bias are computed as 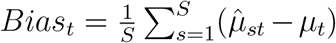 and 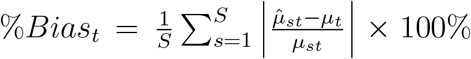 respectively, where *S* = 500 is the number of generated realizations, *µ*_*t*_ is the target value for the mean epidemic curve (step 1 of the simulation in Section 3) and 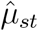 is the corresponding mean nowcast estimate at time *t* and simulation *s*. The prediction interval coverage is obtained by determining the percentage of (true) unreported cases that fall within the interval. Moreover, for each simulation, the interval width is obtained, which is the difference between the upper and lower bound of the prediction interval.

In the simulation, we fix the nowcast date at the end of the month (from March to November). For each nowcast date, the prediction measures are computed for the dates having unreported cases, that is, for *t* = *T* − (*D* − 1), … , *T* . As we have a maximum delay of 7 days, there will be seven dates (including the nowcast date) that involve the prediction of unreported cases. We only present here the simulation results on the nowcast day, that is, at time *T* . Tables 1-4 provide a summary of the results for the prediction measures on the nowcast date for the different functions being considered.

**Table 1:** Performance measures on the nowcast date for function *f*_1_(*t*): **LPS-NB** – LPS model with a negative binomial distribution for the number of cases; **LPS-P** - LPS with a Poisson distribution for the number of cases; **VDK** - Methodology of Van de Kassteele et al. (2019).

| | 95% PI coverage | | | PI width | | | Bias ($\mu$) | | | % Bias ($\mu$) | | |
| --- | --- | --- | --- | --- | --- | --- | --- | --- | --- | --- | --- | --- |
|  | LPS-NB | LPS-P | VDK | LPS-NB | LPS-P | VDK | LPS-NB | LPS-P | VDK | LPS-NB | LPS-P | VDK |
| March | 95.8 | 78.6 | 99.8 | 48.9 | 32.7 | 74.9 | 1.7 | 3.1 | 4.0 | 32.1 | 47.7 | 70.2 |
| Apr | 97.6 | 86.0 | 99.2 | 20.7 | 15.6 | 28.2 | -0.4 | 0.4 | -1.6 | 23.9 | 30.9 | 32.3 |
| May | 96.6 | 73.0 | 100.0 | 41.5 | 25.2 | 73.7 | -1.9 | 1.8 | -3.0 | 19.3 | 26.4 | 27.9 |
| June | 96.0 | 54.0 | 99.6 | 98.6 | 42.2 | 226.5 | -0.1 | 9.3 | 15.3 | 14.7 | 27.7 | 35.3 |
| July | 96.0 | 60.6 | 98.4 | 63.5 | 29.2 | 192.2 | 1.1 | 5.0 | 20.2 | 14.1 | 25.2 | 66.9 |
| Aug | 95.6 | 83.8 | 98.8 | 19.0 | 13.4 | 31.8 | -0.7 | 0.2 | -1.3 | 16.2 | 19.3 | 28.3 |
| Sept | 94.0 | 85.6 | 98.2 | 14.5 | 11.3 | 23.9 | -1.3 | -0.5 | -1.9 | 19.2 | 19.1 | 31.9 |
| Oct | 95.6 | 72.6 | 99.2 | 38.6 | 21.3 | 69.2 | -2.2 | 0.5 | -3.0 | 14.8 | 16.8 | 23.7 |
| Nov | 97.2 | 48.8 | 99.0 | 96.6 | 34.0 | 232.5 | 4.7 | 4.9 | 21.7 | 14.5 | 17.8 | 43.0 |

In terms of prediction interval, the Poisson model (LPS-P) generally has the narrowest prediction interval widths resulting in lower coverage rates for all functions. This is expected since we simulate the data from the negative binomial distribution with an overdispersion parameter which is not accounted for by the Poisson distribution. For function *f*_1_(*t*) (Table 1), the LPS-NB has more stable coverage rates and is closer to the nominal 95% prediction interval across all nowcast dates compared to VDK. The method of VDK on the other hand, tends to have high coverage rates resulting from wider prediction intervals, indicating higher uncertainty in its predictions. For functions *h*_1_(*t*) (Table 2) the PI coverage for LPS-NB and VDK are somewhat similar. The PI width of LPS-NB are wider from March to June and narrower from July to November compared to VDK.

**Table 2:**
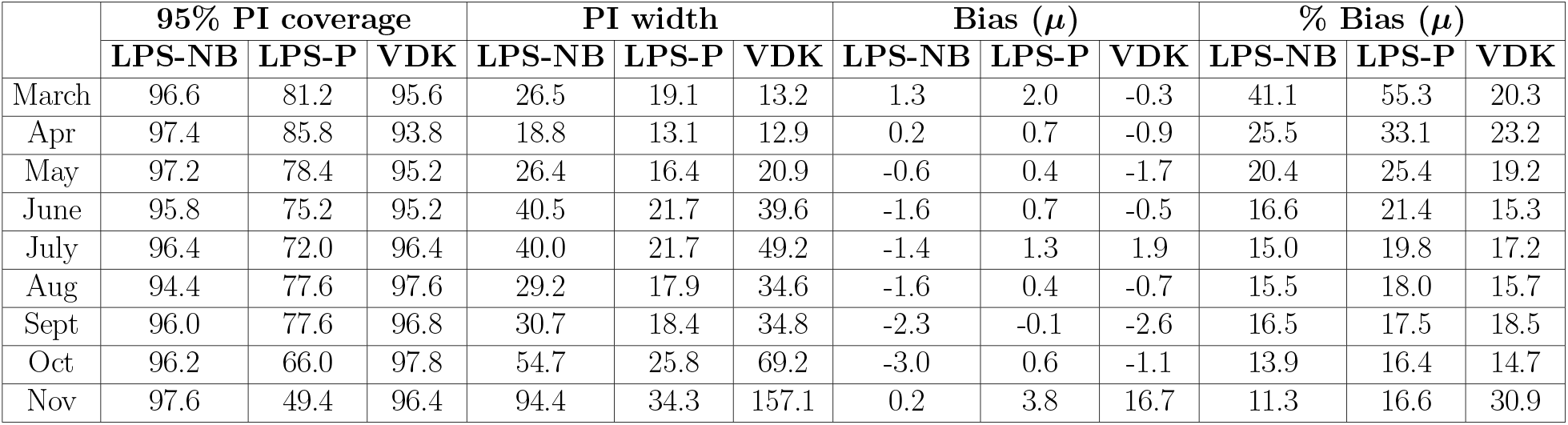
Performance measures on the nowcast date for function *h*_1_(*t*): **LPS-NB** - LPS model with a negative binomial distribution for the number of cases; **LPS-P** - LPS with a Poisson distribution for the number of cases; **VDK** - Methodology of Van de Kassteele et al. (2019).

| | 95% PI coverage | | | PI width | | | Bias ($\mu$) | | | % Bias ($\mu$) | | |
| --- | --- | --- | --- | --- | --- | --- | --- | --- | --- | --- | --- | --- |
|  | LPS-NB | LPS-P | VDK | LPS-NB | LPS-P | VDK | LPS-NB | LPS-P | VDK | LPS-NB | LPS-P | VDK |
| March | 96.6 | 81.2 | 95.6 | 26.5 | 19.1 | 13.2 | 1.3 | 2.0 | -0.3 | 41.1 | 55.3 | 20.3 |
| Apr | 97.4 | 85.8 | 93.8 | 18.8 | 13.1 | 12.9 | 0.2 | 0.7 | -0.9 | 25.5 | 33.1 | 23.2 |
| May | 97.2 | 78.4 | 95.2 | 26.4 | 16.4 | 20.9 | -0.6 | 0.4 | -1.7 | 20.4 | 25.4 | 19.2 |
| June | 95.8 | 75.2 | 95.2 | 40.5 | 21.7 | 39.6 | -1.6 | 0.7 | -0.5 | 16.6 | 21.4 | 15.3 |
| July | 96.4 | 72.0 | 96.4 | 40.0 | 21.7 | 49.2 | -1.4 | 1.3 | 1.9 | 15.0 | 19.8 | 17.2 |
| Aug | 94.4 | 77.6 | 97.6 | 29.2 | 17.9 | 34.6 | -1.6 | 0.4 | -0.7 | 15.5 | 18.0 | 15.7 |
| Sept | 96.0 | 77.6 | 96.8 | 30.7 | 18.4 | 34.8 | -2.3 | -0.1 | -2.6 | 16.5 | 17.5 | 18.5 |
| Oct | 96.2 | 66.0 | 97.8 | 54.7 | 25.8 | 69.2 | -3.0 | 0.6 | -1.1 | 13.9 | 16.4 | 14.7 |
| Nov | 97.6 | 49.4 | 96.4 | 94.4 | 34.3 | 157.1 | 0.2 | 3.8 | 16.7 | 11.3 | 16.6 | 30.9 |

The bias of the three methods varies across functions and months. LPS-NB and LPS-P often exhibit similar bias patterns, while VDK occasionally shows higher biases, particularly for function *f*_1_(*t*) during months like July and November. However, the biases are relatively small in magnitude. In terms of relative bias, VDK sometimes displays higher values compared to LPS-NB and LPS-P, especially noticeable in function *f*_1_(*t*) during months like July and November. For functions *f*_2_(*t*) and *h*_2_(*t*) (Tables 3 and 4) with relatively high case numbers, we did not present the method of Van de Kassteele et al. (2019) since their results yield very wide prediction intervals and large relative bias. We believe that the reason for this is more of a computational issue rather than a methodological one. The LPS-NB model for these functions yield stable results with higher coverage rates, wider PI width, lower relative bias and generally lower bias compared to the LPS-P model.

**Table 3:** Performance measures on the nowcast date for function *f*_2_(*t*): **LPS-NB** - LPS model with a negative binomial distribution for the number of cases; **LPS-P** - LPS with a Poisson distribution for the number of cases.

| | 95% PI coverage | | PI width | | Bias ($\mu$) | | % Bias ($\mu$) | |
| --- | --- | --- | --- | --- | --- | --- | --- | --- |
|  | LPS-NB | LPS-P | LPS-NB | LPS-P | LPS-NB | LPS-P | LPS-NB | LPS-P |
| March | 96.8 | 49.4 | 296.2 | 147.5 | 12.5 | 32.9 | 30.0 | 71.6 |
| Apr | 98.0 | 54.8 | 147.8 | 72.6 | 2.0 | 17.4 | 19.5 | 47.1 |
| May | 98.4 | 50.8 | 142.2 | 63.7 | -3.7 | 14.3 | 16.6 | 32.1 |
| June | 97.4 | 37.0 | 380.4 | 106.5 | 12.2 | 42.5 | 15.7 | 34.5 |
| July | 97.8 | 42.4 | 212.8 | 59.1 | 8.6 | 11.3 | 14.7 | 24.4 |
| Aug | 95.4 | 55.6 | 94.1 | 37.2 | -3.5 | 3.6 | 14.0 | 19.8 |
| Sept | 96.6 | 55.8 | 90.8 | 35.2 | -1.6 | 2.9 | 13.3 | 19.7 |
| Oct | 96.6 | 48.8 | 123.8 | 41.6 | -5.6 | 0.7 | 13.0 | 17.4 |
| Nov | 97.8 | 36.6 | 373.4 | 71.2 | 28.7 | 14.4 | 17.7 | 19.8 |

**Table 4:** Performance measures on the nowcast date for function *h*_2_(*t*): **LPS-NB** - LPS model with a negative binomial distribution for the number of cases; **LPS-P** - LPS with a Poisson distribution for the number of cases.

| | 95% PI coverage | | PI width | | Bias ($\mu$) | | % Bias ($\mu$) | |
| --- | --- | --- | --- | --- | --- | --- | --- | --- |
|  | LPS-NB | LPS-P | LPS-NB | LPS-P | LPS-NB | LPS-P | LPS-NB | LPS-P |
| March | 95.0 | 63.4 | 115.3 | 57.5 | 6.1 | 8.8 | 35.6 | 52.2 |
| Apr | 96.8 | 62.4 | 67.6 | 34.9 | -0.3 | 4.1 | 20.0 | 32.9 |
| May | 97.6 | 53.8 | 103.6 | 45.6 | -2.3 | 7.9 | 16.2 | 27.7 |
| June | 98.0 | 44.4 | 178.6 | 62.0 | -1.1 | 16.8 | 14.0 | 28.3 |
| July | 97.0 | 43.8 | 184.6 | 56.6 | 2.2 | 12.6 | 13.5 | 24.0 |
| Aug | 97.2 | 49.0 | 129.2 | 45.0 | -2.4 | 7.9 | 13.0 | 21.4 |
| Sept | 95.8 | 44.4 | 136.3 | 45.5 | -4.6 | 6.2 | 12.5 | 20.4 |
| Oct | 97.6 | 38.2 | 257.7 | 63.0 | -0.7 | 10.5 | 11.5 | 20.4 |
| Nov | 97.2 | 29.4 | 455.9 | 81.7 | 21.2 | 17.6 | 12.8 | 20.6 |

## 4 Real Data application

We apply our method to COVID-19 mortality data in Belgium for 2021 and incidence data for 2022. The raw data comes from the website of the Sciensano research institute (https://epistat.sciensano.be/covid/; Accessed December 20, 2022). The data contains the cumulative number of cases, reported up to the day of the file. The file is updated every day, and in this way, the number of cases and reporting delays are obtained. The data is structured in matrix format with the date of death/confirmed case as rows and number of days of reporting delay as columns. Figure 4 shows the total number of cases with (dashed) vertical lines corresponding to different nowcast dates used for illustration. As no cases are reported immediately on the day of death/confirmed case in the data, all cases have a delay of at least one day. The proportion of cases reported for each delay is shown in Table 5. Majority of the cases (66% for mortality and 61% for incidence) were reported with a delay of 2 days. Followed by a delay of 1 day (14%) for mortality and 3 days (37%) for incidence. The rest of the delays account for very small percentages of the cases. The reporting delay is truncated to a maximum number of seven days.

**Table 5:** Proportion of cases reported for each delay in the mortality data.

| Delay (days) | 0 | 1 | 2 | 3 | 4 | 5 | 6 | 7 |
| --- | --- | --- | --- | --- | --- | --- | --- | --- |
| Proportion | 0.000 | 0.143 | 0.664 | 0.118 | 0.047 | 0.017 | 0.007 | 0.004 |

**Table 6:** Proportion of cases reported for each delay in the incidence data.

| Delay (days) | 0 | 1 | 2 | 3 | 4 | 5 | 6 | 7 |
| --- | --- | --- | --- | --- | --- | --- | --- | --- |
| Proportion | 0.000 | 0.001 | 0.61 | 0.373 | 0.012 | 0.002 | 0.001 | 0.001 |

**Figure 4.**
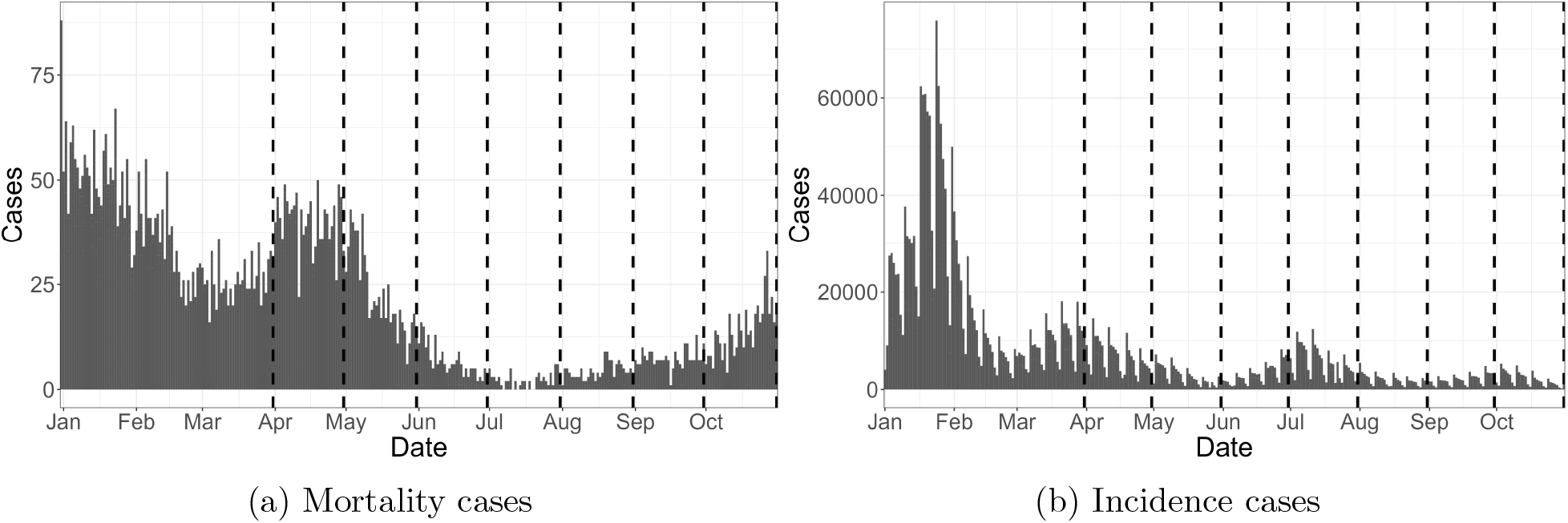
Covid-19 death (a) and incidence (b) cases in Belgium. Dashed vertical lines correspond to different nowcast dates.

We choose *K*_*T*_ = 40 and *K*_*D*_ = 10 for the real data analysis. In addition, we also consider the day of the week effect as an additional covariate in the model. Specifically, we fit the following model log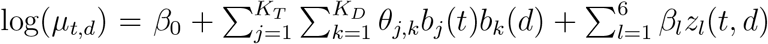 as in (1), where 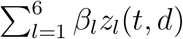 represents the day of the week with Monday taken as the reference category. Algorithms to fit the model are available within the EpiLPS package (Gressani, 2021) through the *nowcasting()* routine.

Figure 5 presents the nowcasting results using the mortality data at different nowcast dates (namely, at every end of the month). The blue color represents the reported cases for the past 14 days on the nowcast date, the gray color represents cases that have not yet been reported, and the red points with error bars corresponds to the nowcast prediction with the 95% prediction interval. It can be seen that most of the nowcast predictions on the nowcast date are fairly close to the observed cases (gray). In addition, all of the observed cases fall within the prediction interval. For the incidence data, we used a maximum delay of 5 days. The nowcast results for the incidence data (Figure 6) tend to exhibit varying degrees of accuracy. This means that some nowcast values are close to the observed values, while others are further away, and some are moderately close. Notably, the nowcast estimate (on the nowcast day) for May and July are somewhat distant from the observed values.

**Figure 5.**
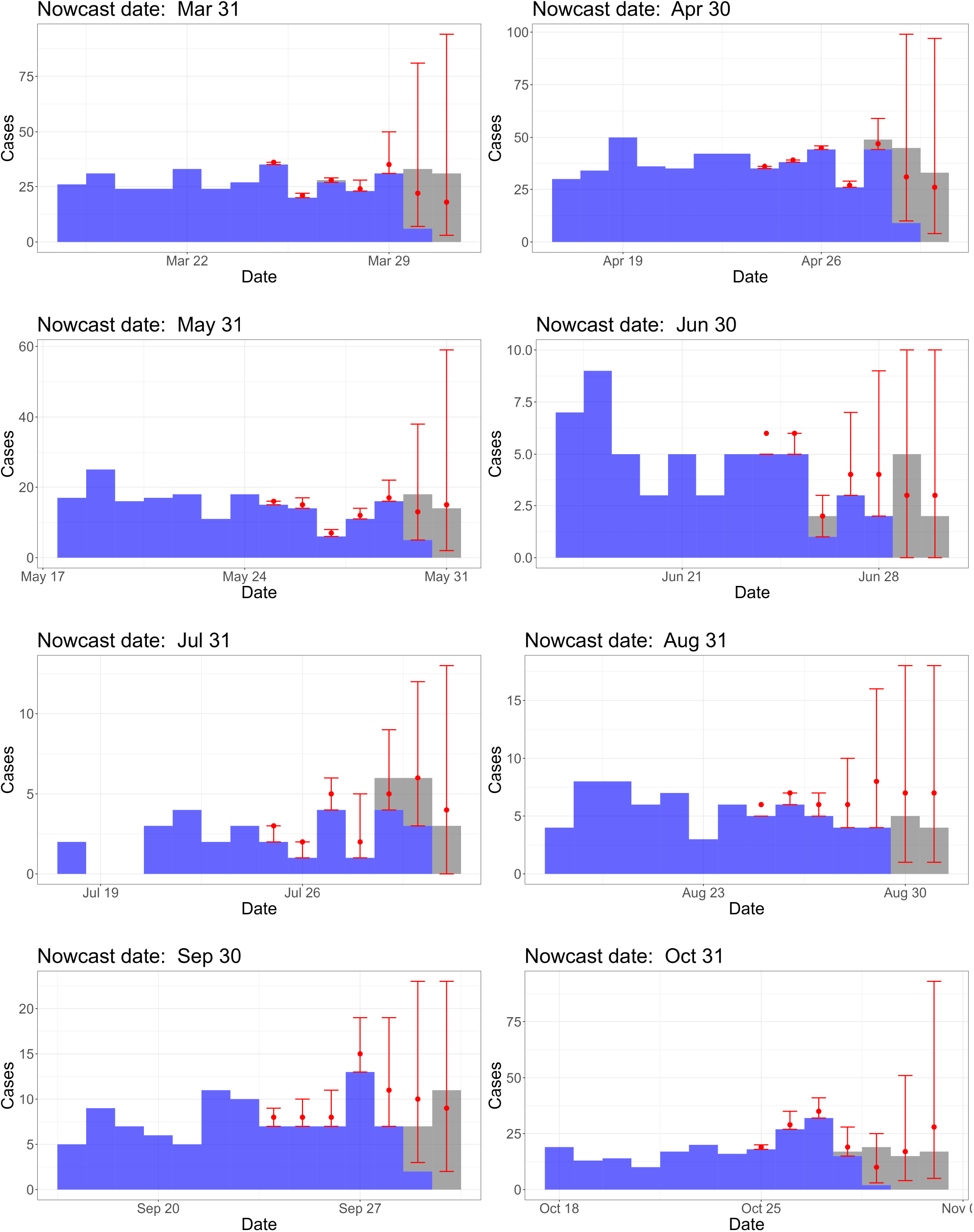
Nowcast for mortality data with different nowcast dates. Blue - reported cases ; Gray - not-yet-reported cases; Red points - nowcast estimates; Red error bar - 95% nowcast prediction interval.

**Figure 6.**
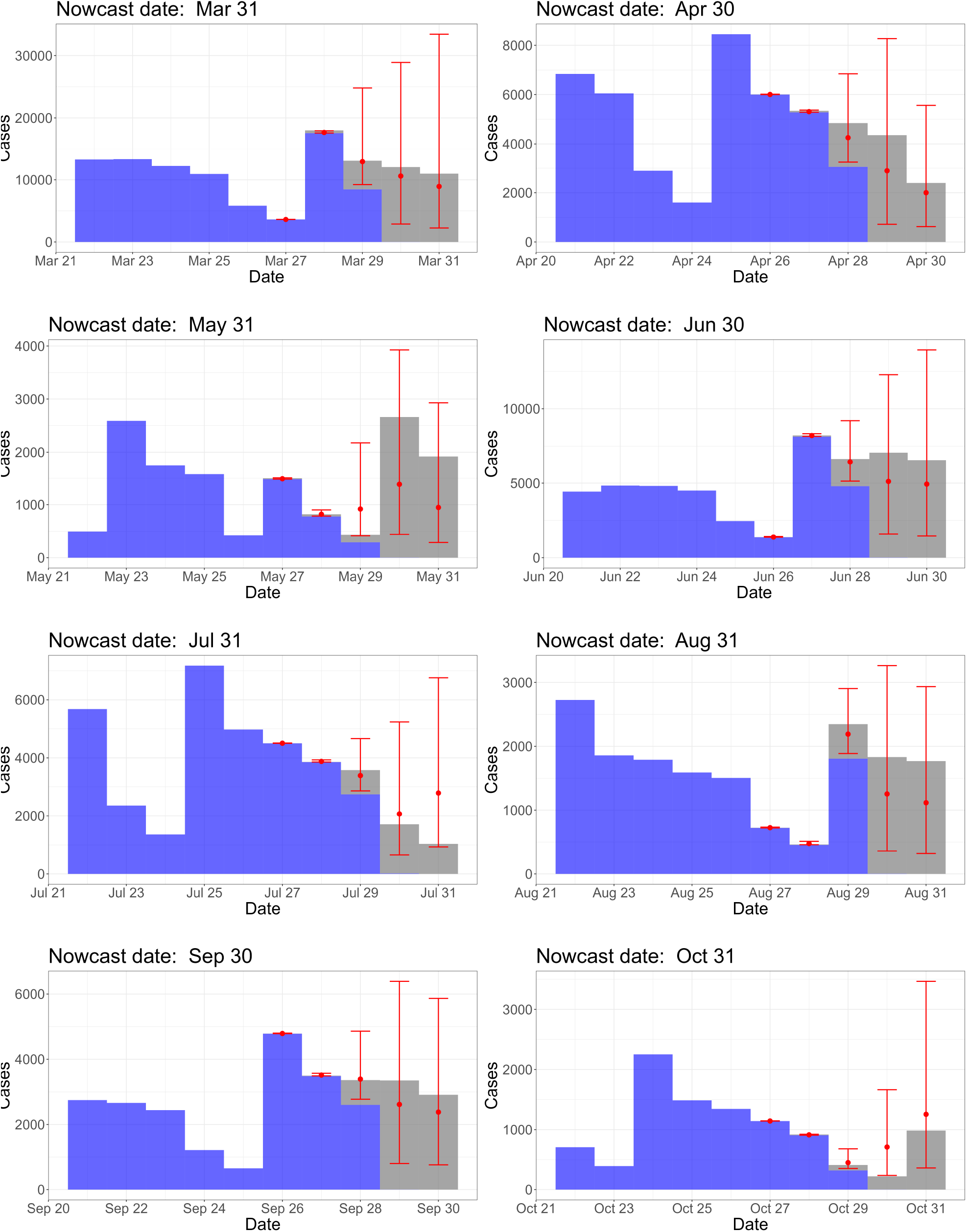
Nowcast for incidence data with different nowcast dates. Blue - reported cases ; Gray - not-yet-reported cases; Red points - nowcast estimates; Red error bar - 95% nowcast prediction interval.

The plot for the estimated delay density is shown in the supplementary material S5. The steps to obtain the time-varying delay distribution is outlined in Section 2.5. The plots show that the delay distribution is fairly constant through time for both incidence and mortality data. The density is highest for the delay of 2 days except in the beginning of the year for incidence data. This confirms the observed reporting intensity in our data that most cases are reported with a two-day delay.

## 5 Conclusion

This paper presents a novel Bayesian approach of the nowcasting model proposed by Van de Kassteele et al. (2019). During a pandemic or outbreak, it is very important to get an idea on the number of actual cases in real-time. One advantage of using the Bayesian framework is that prediction intervals can be naturally obtained. Our proposed methodology is based on a combination of Laplace approximations and P-splines, making it both fast and flexible.

Based on the results presented in this paper, Bayesian nowcasting with Laplacian-P-Splines seems to be a promising tool. We performed a simulation study to evaluate the (predictive) performance of our method. Simulation results demonstrate that our proposed method, under the assumption of a negative binomial distribution, consistently produces stable results with excellent prediction interval coverage that is closer to the nominal level and exhibits small bias. Moreover, we also applied our method to the COVID-19 mortality and incidence data in Belgium. The nowcast predictions for the mortality data seem to be fairly close to the actual observed values, and all of the 95% prediction intervals for the different nowcast dates being considered contained the observed values. In the case of incidence data with larger numbers of cases, the nowcast results tend to exhibit varying degrees of accuracy. However, it is important to take note that we have no data for the nowcast day, and there are very few reported cases (0.1%) with a one-day delay for the incidence data. This makes it much more difficult to produce accurate nowcast predictions.

While nowcasting can provide valuable real-time information and predictions, it also has certain drawbacks. Nowcasting heavily relies on real-time data, which may not always be readily available, or of good quality. Biases in the data (such as double-counting of cases) that are corrected at a later time, can impact the nowcasting prediction. In addition, gaps in data collection, processing, or dissemination can impact the timeliness and effectiveness of nowcasting predictions. Despite these drawbacks, nowcasting has been a very important tool for obtaining timely information and short-term predictions. While the proposed nowcasting model is rather complex, we have coded the *nowcasting()* routine in the EpiLPS package (Gressani (2021)) to provide a user-friendly experience. Finally, one extension to the LPS methodology for nowcasting is to incorporate the now-casting model into the estimation of the reproduction number in the method proposed by Gressani et al. (2022b). This has been currently implemented by Sumalinab et al. (2023). Another extension is to consider the correlation of cases for each time point (*t*). By doing this, the model can capture temporal dependencies and may provide better estimates of the variability. Furthermore, accounting for spatial correlation and other covariate effects would be valuable. Incorporating spatial information can account for local variations while considering additional covariates can provide a better understanding of the factors influencing disease spread.

## Data Availability

The raw data is available on the website of the Sciensano research institute at https://epistat.sciensano.be/covid/covid19_historicaldata.html.

https://github.com/bryansumalinab/Laplacian-P-spline-nowcasting.git

https://epistat.sciensano.be/covid/covid19_historicaldata.html

## Supplementary materials

**Online Appendix**: Contains details for the model matrices and parameters, derivations for the gradient and Hessian, hyperparameter optimizations, details for the LPS-Poisson model, supplementary figures, and tables. (supplement.pdf)

**R scripts**: The R codes used to implement the simulation study are available on GitHub through the link https://github.com/bryansumalinab/Laplacian-P-spline-nowcasting.git.

